# International Adaptation of a brief Problem-Solving Skills (the IAPPS trial) training for people in custody with severe mental illness in Poland: an open multicentred, parallel group, feasibility randomised controlled trial

**DOI:** 10.64898/2026.04.24.26351654

**Authors:** Amanda E Perry, Maja Zawadzka, Jaroslaw Rychlik, Catherine Hewitt, Sarah Morris

## Abstract

**Objectives:** The primary aim of this study was to assess the feasibility of delivering an adapted problem-solving skills (PSS) intervention by quantifying the recruitment, follow-up and completion rates using a brief problem-solving intervention for people with a mental health diagnosis in two Polish prisons.

**Design:** IAPPS is an open, multi-centred, parallel group feasibility randomised controlled trial (RCT).

**Setting:** Two prisons in Poland.

**Participants:** Men in custody aged 18 years and older, having a mental illness and living within the prison therapeutic unit.

**Interventions:** The intervention consisted of an adapted PSS skills intervention plus care as usual (CAU) or care as usual only. Delivered in groups of up to five people in 1.5-hour sessions over the course of two weeks.

**Main outcome measures:** Primary outcomes - rate of recruitment, follow-up, and feasibility to deliver the intervention. Secondary outcomes included measures of depression, general mental health, and coping strategies.

**Results:** 129 male prisoners were screened, 64 were randomly allocated, with a mean age of 53.5 years (SD 14, range 23-84). 59 (95%) prisoners were of Polish origin. Our recruitment rate was 48%. There was differential follow up with those in the intervention group less likely to complete the post-test battery versus those who received care as usual. Outcome measures were successfully collected at both time points.

**Conclusions:** We were able to recruit, retain and deliver the intervention within the prison setting; some logistical challenges limited our assessment of intervention engagement. Our data helps to demonstrate how use of the RCT study design can be implemented and delivered within the complex prison environment.

**Trial registration number:** ISRCTN 70138247, protocol registration date May 2021 (insert link).

**STRENGTHS AND LIMITATIONS OF THIS STUDY:** This is the first feasibility RCT to examine the recruitment, follow-up, and completion of measures to a brief PSS training for adult males with mental health problems in two Polish prisons. Results showed promising recruitment rates and retention to the intervention delivery. Whilst the study was not designed to test effectiveness, we demonstrated importantly that delivering brief interventions for people in custody were feasible and acceptable.

## INTRODUCTION

The mental health of people incarcerated in prison is recognized as a worldwide public health concern.^1-3^ People residing in prison experience a disproportionate level of mental health problems, (particularly major depression,^3-5^ self-harm and anti-social violent behaviour) than in the general population.^3,6-8^ Isolation and boredom within prisons are linked to poor mental health and can lead to an increased likelihood of problems becoming exacerbated.^9^ Internationally over the last five years, prisons have reported an unprecedented rise in the incidence of violent assaults and suicidal behaviours,^10,11^ and the co-morbidity between these elements are well documented.^5,8,12,13^

A small proportion of people in prison receive some kind of support, but generally the demand outstrips the available resources. PSS is a brief intervention based upon cognitive behavioural principles has been extensively tested with several patient groups in the community^14-17^. Trials of PSS report reductions in depression and allied constructs such as hopelessness.^16,18,19^ Such skills can be delivered by different professional groups and lay persons, and the World Health Organization (WHO) has adopted the general model of ‘problem management’ to help those dealing with international crisis situations. ^17,18^ The link between use of problem-solving skills and symptoms of depression, self-harm and/or violent behaviour is acknowledged in the literature as many cite the immediate cause of such behaviour as a problem or recognised problems in their lives.^14,20,21^ For these reasons, the simplicity of the skills, and the brief nature of the delivery mode of delivery lends itself to supporting people in custody where they experience complex problems and resources for individualised therapy are scarce.

Previous UK studies have shown promising results for the use of problem-solving skills in prisons ^22-25^ and accredited Her Majesty Prison and Probation Service courses (Enhanced Thinking Skills) have been used and evaluated in UK prisons for several years^26^. Although other European countries have not yet used such techniques;^27^ the necessary cultural and environmental adaptations are necessary to ensure good adherence and acceptance of those that are receiving it. In our prior work, we adapted a UK prison-based PSS model for use in Polish prisons.^28^ Other cultures acknowledge the challenges relating to the disclosure of mental health problems and for Polish people in custody the reluctance of having to admit to having a mental health problem promotes little opportunity for social interaction or intervention delivery.^27^ This study was used to evaluate the feasibility, acceptability, and delivery of the adapted UK prison-based PSS model ^22,24^ with people with a mental health diagnosis incarcerated in two Polish prisons.

## METHOD

### Study design

The trial protocol (ISRCTN70138247; see Materials)^29^ describes an open, 12-month parallel group, feasibility RCT with follow-up assessments at ten weeks after randomisation. Participants were recruited from two ‘closed prison facilities with a therapeutic wing’ in Poland (ZK Racibórz, and ZK Kłodzko). Together the prisons holding up to 1,474 male adults representing people on remand, pre-trial detainees, first time offenders and non-psychotic prisoners with a mental health diagnosis. The trial received ethical approval from the Department of Health Sciences; University of York; Research Governance Committee on 13^th^ May 2021; from the Governor at each prison site and the Polish equivalent of the GDPR expert (RODO) within Poland.^29^ Eligible participants included male adults living on the prison therapeutic wing (18 years or older), having a sentence length or planned duration of more than three months. Exclusion criteria included anyone who was unable to provide informed consent, had a learning disability or were considered by the prison staff to pose a risk to the research team.

### Recruitment

All participants residing on the therapeutic wing of each prison were approached individually by the research team (MZ and JR) to assess their willingness to participate in the study over a four to six - day period in July 2021. Any concerns about participant ill health (either physically or mentally) resulted in referral to the prison clinical team (i.e., doctor or psychologist) as per the prison protocol. After an independent examination by the clinical team, decisions were made about whether the participant was well enough to participate in the study. Following the initial approach, those willing to learn more about the study were invited to attend a group meeting for up to one hour with up to five other participants. The meeting described the purpose of the study and gave an opportunity for participants to ask any questions. Participants had the right to decline and were able to leave the meeting if they did not want to take part. Willing participants received a copy of the Participant Information Sheet (PIS: insert link), privacy notice and consent form; all were read verbatim to the group. A written informed consent was followed by completion of the baseline assessment and a demographic questionnaire. Participants willing to take part in the study were given a unique identification study number used to anonymise individual participant data. An anonymised secure, encrypted datasheet was shared across the organisations. This also blind the statistician to the allocation of the groups.

### Randomisation Procedure

The randomisation sequence was generated by a statistician in York Trials Unit at the University of York in the UK using stratified block randomisation with stratification by prison site and with varying block sizes of 4 and 6. Access to the sequence code was confined to the Trial Manager. Following baseline assessments each participant and member of the research team were made aware of their treatment allocation via the secure database.

### The problem-solving skills intervention

The brief PSS intervention involved delivery of an adapted UK problem-solving training package^28^ that used established social problem-solving theory.^,30-32^ This comprised of a seven-step problem-solving model comprised of: Step one: ‘is there a problem’? Step two: ‘describe the problem’; Step three: getting information; Step four: ‘think of options’; Step five: ‘choose an option’; and Step six: ‘make a plan’. Delivery of the intervention was conducted by a qualified Cognitive Behavioural Psychotherapist (MZ) in groups of up to five participants, with each session lasting up to 1.5 hours over the course of two weeks. The intervention delivery comprised of a five-minute digital animation which presented a case study of someone in custody talking about how they managed to navigate a relationship breakdown, completion of a workbook and demonstration of the problem-solving skills by MZ. At the end of the session participants were asked to practice the skills and finish the workbook. All participants received a notebook and calendar as a token of appreciation. All participants regardless of their allocation received care as usual. Care as usual comprised of any service and or treatment provided by prison staff and/or medical staff as per the prison and healthcare protocols.

### Outcome Measures

A demographic questionnaire collected participant self-report information on their mental health, criminal activity, and protected characteristics. Primary outcomes included the recruitment rate, follow-up and feasibility as determined by rates of attrition. At baseline and follow-up, the assessment comprised of three secondary outcome measures including depression (PHQ-9^33-35^), general mental health (GHQ-28,^36,37^) and coping strategies (COPE-36^38.^). Adverse events were reported using a standard protocol as per the existing arrangements from within the prison.

### Sample size

As this was a feasibility trial, the sample size was based on estimating recruitment and follow-up rates. A recruitment target of up to 100 randomised participants with an estimation of recruitment (50%) and follow up rates (80%) to be estimated within a 7% and 8% margin of error.^35^ The research was not designed or powered to detect a statistically significant difference between time points.

### Statistical analysis

Data were analysed using IBM SPSS Statistics. The primary outcome: recruitment rate (defined as the number of recruited participants divided by the number of eligible participants), was estimated and presented along with a 95% confidence interval. The questionnaire retention rate (number of returned questionnaires divided by the number of randomised participants) at 10 weeks post-randomisation was calculated. The secondary outcome measures (PHQ-9; GHQ-28; COPE-36)^33-38^ were reported descriptively using mean, SD, median, 25th and 75th percentiles for continuous outcomes and the number of events and percentages for categorical data. The number of adverse events were described as per the prison protocol. All missing data was treat as missing and no imputation was applied. All participant data were analysed on an intent to treat basis.

### Patient and public involvement

This study was part of a larger phase of work which included extensive work to adapt the intervention for use within polish prisons.^28.^The results of this study directly informed the study design and the materials that were used to deliver the training. No specific patient and public involvement activities were conducted for this study.

## RESULTS

The flow of participants through the trial was detailed in a CONSORT diagram,^41^ (Figure 1). The number of participants enrolled, randomly allocated, completing follow up and analysed is presented alongside recorded reasons withdrawal from the study. Between the months of July 2021 and August 2021, 129 participants were assessed for eligibility. A high proportion were excluded, (n=67); 58 declined to participate, 5 refused to take part after consent but prior to randomisation, and 2 individuals were deemed a risk to the research team. The remaining 64 participants consented and were randomised to either PSS and care as usual (n=31) or care as usual alone(n=33).

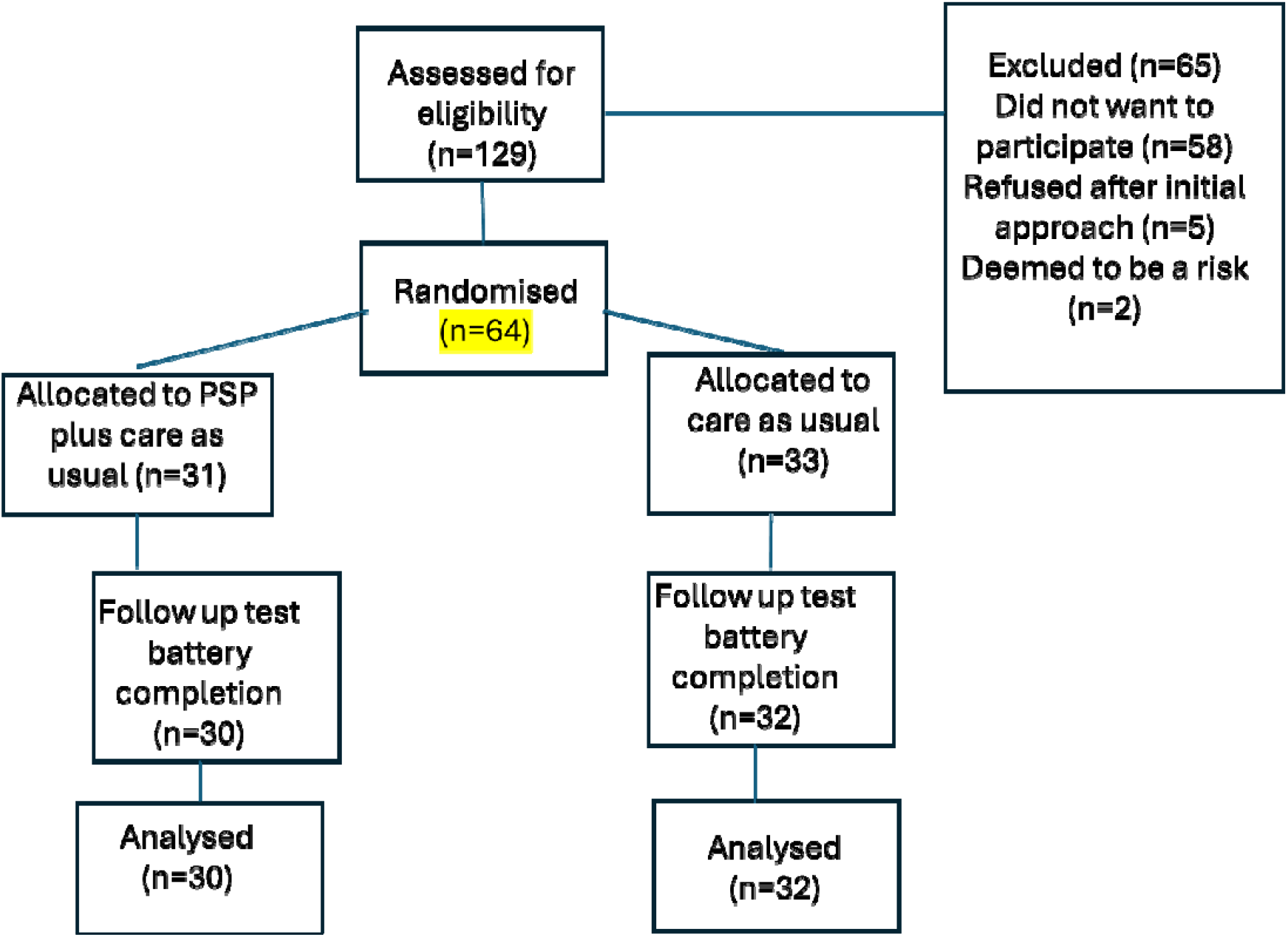

Participants’ mean age was 53.5 years (range 23 to 84 years). Most of the participants were Polish (59, 95%), single (46, 75%) and reported it was not their first time in prison (53, 85%). Most participants had been in prison between 1 and 5 times and on average had served 136 months. All had a life sentence and over half recognised that problems in their lives had impacted on their mental health (see Table 1).

**Table 1:**
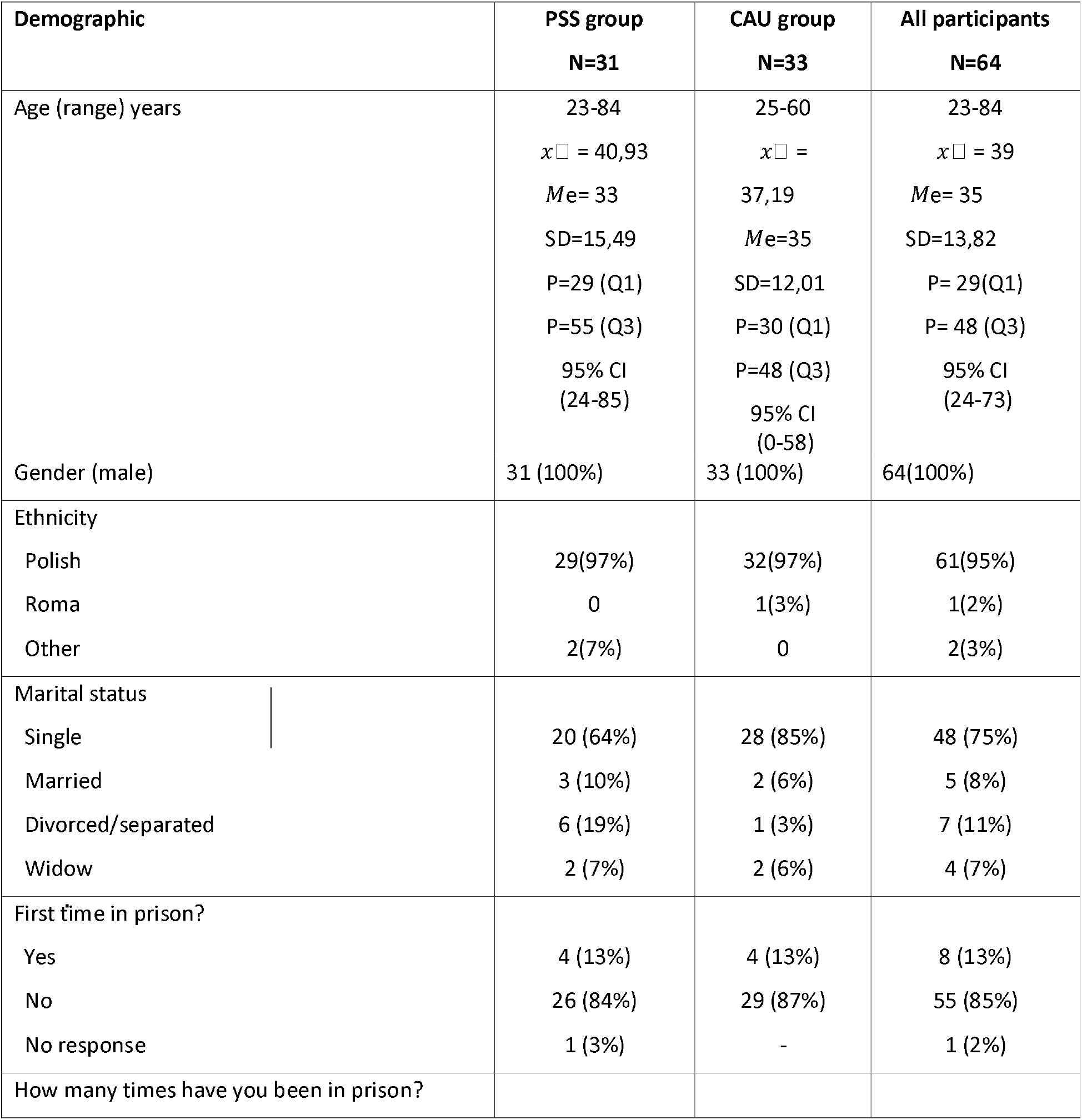
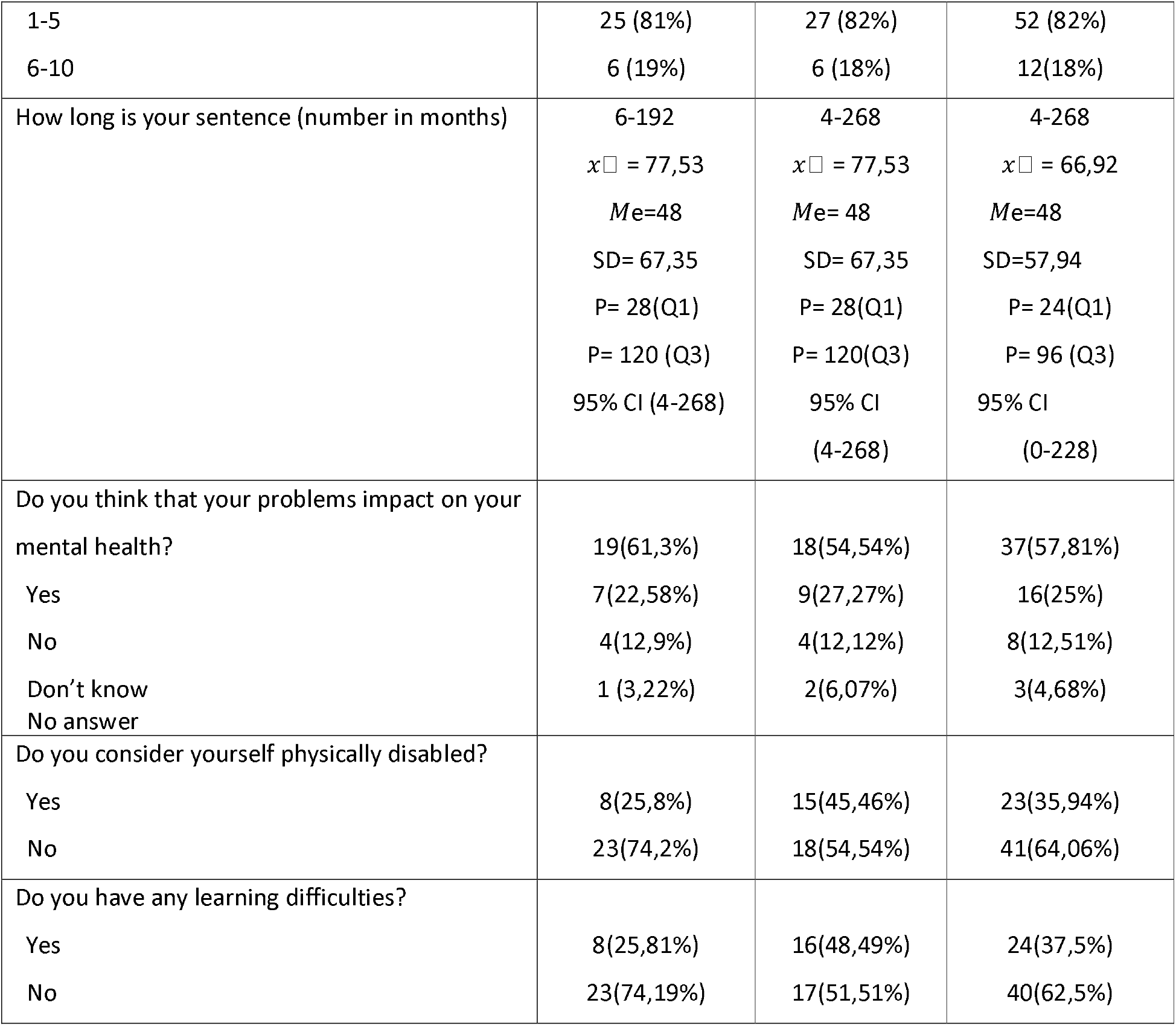
Sociodemographic characteristics at baseline.

### Primary outcomes

The recruitment took place at both prison sites at a similar rate, taking six (in Raciborz) and five days (in Klodzko) during July and August 2021. The overall recruitment rate (RR: offers accepted/offers made *100) was 48%. Retention in completion of post-test outcomes was 96% at ten weeks post intervention: PSS and CAU, (n=30); vs CAU, (n=32). Feasibility to deliver the intervention was affected by the prison regime. Delivery of the intervention was condensed into one session and post-treatment data were collected at ten not six weeks.

### Secondary outcomes

Secondary outcomes collected at 10 weeks post-randomisation (see Table 2). At post-test scores were slightly lower for depression and social dysfunction in the intervention group compared to the usual care group. Scores post-test on coping outcomes reported via the COPE-36 were similar within both groups.

**Table 2:**
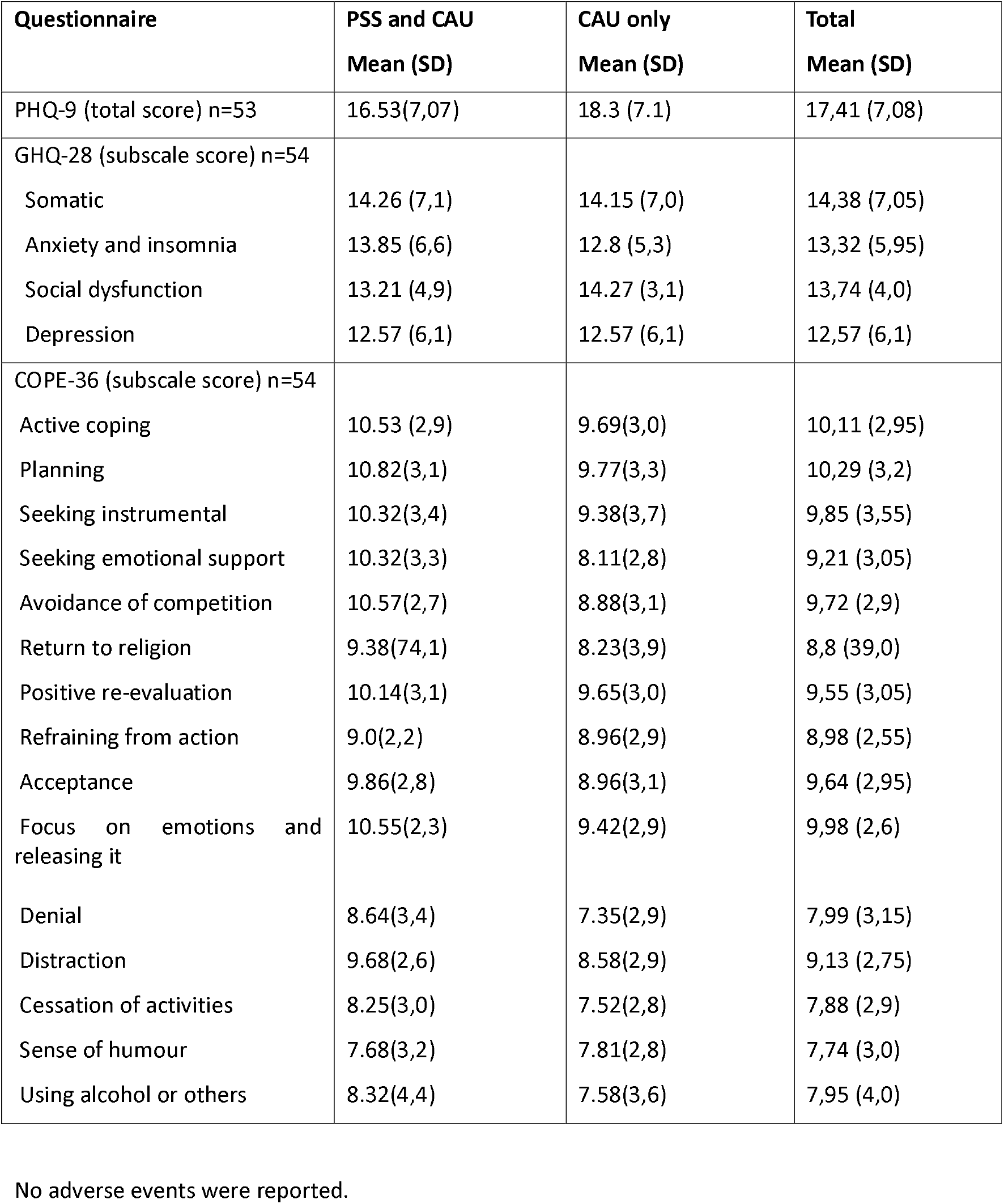
Secondary outcome measures – post-test follow-up.

| Questionnaire | PSS and CAU<br>Mean (SD) | CAU only<br>Mean (SD) | Total<br>Mean (SD) |
| --- | --- | --- | --- |
| PHQ-9 (total score) n=53 | 16.53(7,07) | 18.3 (7.1) | 17,41 (7,08) |
| GHQ-28 (subscale score) n=54 |  |  |  |
| Somatic | 14.26 (7,1) | 14.15 (7,0) | 14,38 (7,05) |
| Anxiety and insomnia | 13.85 (6,6) | 12.8 (5,3) | 13,32 (5,95) |
| Social dysfunction | 13.21 (4,9) | 14.27 (3,1) | 13,74 (4,0) |
| Depression | 12.57 (6,1) | 12.57 (6,1) | 12,57 (6,1) |
| COPE-36 (subscale score) n=54 |  |  |  |
| Active coping | 10.53 (2,9) | 9.69(3,0) | 10,11 (2,95) |
| Planning | 10.82(3,1) | 9.77(3,3) | 10,29 (3,2) |
| Seeking instrumental | 10.32(3,4) | 9.38(3,7) | 9,85 (3,55) |
| Seeking emotional support | 10.32(3,3) | 8.11(2,8) | 9,21 (3,05) |
| Avoidance of competition | 10.57(2,7) | 8.88(3,1) | 9,72 (2,9) |
| Return to religion | 9.38(74,1) | 8.23(3,9) | 8,8 (39,0) |
| Positive re-evaluation | 10.14(3,1) | 9.65(3,0) | 9,55 (3,05) |
| Refraining from action | 9.0(2,2) | 8.96(2,9) | 8,98 (2,55) |
| Acceptance | 9.86(2,8) | 8.96(3,1) | 9,64 (2,95) |
| Focus on emotions and releasing it | 10.55(2,3) | 9.42(2,9) | 9,98 (2,6) |
| Denial | 8.64(3,4) | 7.35(2,9) | 7,99 (3,15) |
| Distraction | 9.68(2,6) | 8.58(2,9) | 9,13 (2,75) |
| Cessation of activities | 8.25(3,0) | 7.52(2,8) | 7,88 (2,9) |
| Sense of humour | 7.68(3,2) | 7.81(2,8) | 7,74 (3,0) |
| Using alcohol or others | 8.32(4,4) | 7.58(3,6) | 7,95 (4,0) |

No adverse events were reported.

## DISCUSSION

This study is one of the first to establish the recruitment, retention, acceptability, and engagement of a brief PSS training intervention for people in custody with significant mental health problems in Poland. Preliminary evidence suggests that we have been able to assess the feasibility of how to examine this information to inform a definitive large RCT. The numbers randomised fell short of the 80 participants target known to be the established figure for good quality feasibility work^39,40^. Nevertheless, the recruitment and follow-up rates were encouraging at 48% and xx respectively. Conducting research in a complex prison environment is often limited by the constraints of the regime and the unpredictable nature of the space in which researchers are working^42.^ The higher-than-expected post-test completion rate was probably an artefact of this specific population in which most of the group were in prison for life sentences (62/64, 96%) coupled with the nature of the specialist unit in which they lived. For this reason, this population of people in custody is likely to be more stable than those that have the opportunity for release or transfer to another prison allowing them to progress through their sentence.

Other research has shown similar recruitment rates in prisons^43^ demonstrating that with careful planning successful recruitment with this population is possible. The unique nature of the environment, the population and the custodial setting exacerbates participant concerns around the sharing of data not just in UK prisons but as an international concern^44^ coupled with the challenges of motivating people with severe mental illness reported in allied community populations^45.^ This was particularly apparent in our study where the mental health conditions included those with personality disorders, (n=11, 18%); hyperactivity, (n=11, 18%); schizophrenia and or psychotic disorders, (n=10; 16%). The information does, however, help us to estimate the anticipated recruitment time for a larger trial based on the numbers of those that need to be approached. Other trials in the community have shown that recruitment to trials is one of the biggest factors affecting the cost and delivery of the trial; with many requesting extensions to the original scope^46^. The information in this study helps guide that notion of recruitment and the amount of resources required to conduct a large-scale randomised controlled trial in this setting.

### STRENGHTS AND LIMITATIONS

This study has several limitations. We experienced two deviations from the original protocol; first the original post-test measure was to be conducted at six weeks but restrictions within the prison meant that this had to completed at a ten-week follow-up, and second, we had intended to measure acceptability of engagement using several intervention sessions. We delivered one session due to the limitations of the prison regime and access to the study resources. This limited us to fully explore varying levels of engagement over time. The findings are unique to this prison population, furthermore it is unlikely that these findings will be generalisable to other groups of people in custody without complex mental health problems. The results are based on a limited follow-up period (of ten weeks). A longer follow-up may have resulted in some attrition in those completing the test battery. Lastly, no females took part in this study; representation of interventions targeting the gendered needs of all those in custody are an important consideration; and further exploration within this population is required.

## CONCLUSION

We have acknowledged the value of feasibility, resources and understanding more about the time taken to recruit within this population. Such information will inform a pragmatic approach to future research and the design of a larger RCT. It is also worth not noting that no other trials to our knowledge have tested the feasibility of using PSS skills in such a group of people in custody. Conducting this trial in a real-world setting has allowed us to assess the feasibility, acceptability, and adherence to the intervention delivery. In conclusion, this study makes an important contribution in demonstrating how RCTs can be implemented and delivered successfully with individuals who are marginalised and often social excluded from society. Although we acknowledge that further research is necessary to establish intervention effectiveness and generalisability more firmly, this study is an important step in understanding how to conduct high quality research to ultimately improve the lives of people living with mental health problems in custody.

## Supporting information

Trial protocol

## Data Availability

All data produced in the present study are available upon reasonable request to the authors

## Acknowledgements

We acknowledge the support of the prison governors and prison officers at each prison within Poland (ZK Racibórz, and ZK Kłodzko).

## Contributors

AP is the primary author and principal investigator on the study. All authors contributed to the conceptualisation, implementation, analysis & interpretation of data including the literature review, study design, data collection, data analysis, data interpretation, writing. In addition to the above, MZ and JR delivered the intervention and took all baseline and post assessment measures in the two Polish prison sites, statistical advice and support were provided by and reviewed by CH, SM edited and proofed the manuscript. All authors reviewed the manuscript.

## Disclaimer

The views expressed are those of the authors and not necessarily of the Centre for Future Health, nor the Polish penitentiary system.

## Patient and Public involvement

Patients and or public were involved in the development and design of the materials that were developed and adapted for use within this trial. Refer to the methods section for further details.

## Patient consent for publication

Consent obtained directly from the patients

## Ethics approval

The study involves human participants and was approved by the Health Sciences Research Governance Committee on 13^th^ May 2021; from the Governor at each prison site and the Polish equivalent of the GDPR expert (RODO) within Poland. Participants gave informed consent to participate in the study before taking part.

## Provenance and peer review

Not commissioned, externally peer reviewed.

## Data availability statement

Data collected for this study, including de-identified individual participant data is available on reasonable request from the PI. The study protocol is available on request. All requests for data access will need to specify the planned use of data and will require approval from the trial investigator team and the funder prior to release.

## Open access

This is an open access article distributed in accordance with the creative commons attribution licence, which permits others to distribute, remix, adapt, build upon this work non-commercially and licence their derivative works on different terms, provided the original work is properly cited, appropriate credit is given and any changes made indicated and the use is non-commercial.

