## Supplementary material for "International Adaptation of a brief Problem-Solving Skills (the IAPPS trial) training for people in custody with severe mental illness in Poland: an open multicentred, parallel group, feasibility randomised controlled trial": Trial protocol

**Study Protocol version 2 registered on:18/5/2021**

**Published on: 20 /05 /2021**

**Study start date: 15/10/2020**

**Study project end date: 15/10/2021**

**Title: I**nternational **A**daptation of **P**roblem-**S**olving **S**kills in Poland (IAPSS): A study protocol for a feasibility randomized controlled trial for offenders in custody to improve symptoms of depression, general well-being and coping strategies.

**Names protocol contributors**

Dr. Amanda E. Perry, Senior Lecturer, Health Sciences Department, University of York, UK.

Maja Zawadzka, Academic Teacher, The Academy of Justice, Warsaw, Poland.

Dr. Jaroslaw Rychlik, Assistant Professor, The Academy of Justice, Warsaw, Poland.

Professor Catherine Hewitt, Trials Unit, University of York, UK.

**Abstract**

- **Background**: Depression, mental health, and wellbeing are international public health problems affecting large numbers of incarcerated people.
- **Methods**: The study will pilot the use of a brief problem-solving intervention to determine feasibility and acceptability of the proposed recruitment methods, research design and delivery of the intervention training materials. Up to 100 individuals in two Polish prisons will be randomized in a feasibility trial to receive either a problem-solving intervention plus usual care (n=50) or usual care only (n=50). Clinical outcome measures include depression, general mental health and well-being and coping strategies.
- **Discussion**: This study will evaluate the feasibility and acceptability of a brief problem-solving intervention. The study will serve as formative work for a larger evaluation in this population.
- **Trial registration**:
  Published prospectively on 18/05/2021, registration name: [insert ISRCTN NUMBER]. The trial is supported by the York Trials Unit (https://www.york.ac.uk/healthsciences/research/trials/). The project is funded in collaboration with the Centre for Future Health, University of York.

**Keywords:** feasibility, mental health, prisoners, Poland, problem-solving, depression, offenders, randomized controlled trial.

**1.0 Introduction**

The mental health of people incarcerated in prison is recognized as a worldwide public health concern[1-3]. People residing in prison experience a disproportionate level of mental health problems, (particularly major depression[3-5]) self-harm and anti-social violent behavior than in the general population[3,6-8]. Isolation and boredom within prisons are linked to poor mental health and can lead to an increased likelihood of problems becoming exacerbated[9]. In the last five years, UK prisons have reported an unprecedented rise in the incidence of violent assaults and suicidal behaviours;[10,11,] and the co-morbidity between these elements are well documented[5,8,12,13].

Problem-solving therapy (PST) is one example of a psychosocial intervention that has been extensively tested with a range of health patients in the community[14-17]. Trials of PST report reductions in depression and allied constructs such as hopelessness[16,18,19]. Such skills can be delivered by a range of professional groups and lay persons and are used by the World Health Organization to help those dealing with international crisis situations[17,18]. Many people who display symptoms of depression, self-harm and/or violent behavior report the main immediate cause as being problems in their lives[14,20-22]. For these reasons, the simplicity of the skills and their mode of delivery lends itself to support prisoners who often experience complex problems during incarceration. Previous UK studies have shown promising results for the use of problem-solving skills in prisons, but a large-scale evaluation of effectiveness is required[23-26].

Although cognitive behavioral therapy (CBT) approaches have been used as part of the UK prison service accredited programs to help support people who are incarcerated, such techniques have yet to be employed in Polish prisons. Studies have shown that polish prisoners are reluctant to admit having mental health problems and to cooperate with prison staff. An essential prevention element in a prison environment is therefore having a possibility of social interaction or intervention.[27] A wide range of specialist psychotherapeutic interventions should also take into account rehabilitation programs related to the current achievements of clinical practice in the therapy of comorbid mental health problems. In the Polish penitentiary system, the achievements of cognitive-behavioral psychotherapy are primarily used in the treatment of aggressive prisoners and sexual offenders. Consequently, widespread use of brief therapeutic interventions are yet to be explored.[28]

To address this gap within the Polish prison system; the overall aim of the study is to evaluate the acceptability and feasibility of adapting and delivering the UK problem-solving training package[25,26] for use with Polish prisoners.

### 2.0 Aims and objectives

Our overall objective is to assess the feasibility and acceptability of a brief problem-solving intervention and report on clinical outcome measures of depression, general mental health and well-being and coping strategies.

### 2.1 Trial design

The trial design is a proposed multi-centered 2-armed individual randomized controlled feasibility trial with two conditions of treatment (brief problem-solving intervention plus care as usual (CAU) vs control (CAU only).

### 2.2 Methods: Participants, interventions, and outcomes

#### 2.2.1 Study setting

The study will take place in two Polish prisons (zk Racibórz and zk Klodzko). Together the prisons hold up to 1,493 male adult prisoners. Prisoners residing in the prisons represent those on remand, pre-trial detainees and non-psychotic prisoners with a mental health diagnosis who are housed on a specific therapeutic wing. Both prisons are classified as 'semi-open' with a proportion of those serving sentences for the first time.

#### 2.2.3 Eligibility criteria

Adult male prisoners >18 years of age, with a mental health diagnosis and housed on the therapeutic unit. We will exclude prisoners whose: i) length of sentence or planned duration is less than three months, ii) prisoners who are unable to provided informed consent and/or iii) prisoners who pose a risk to the researchers and/or (iv) have a learning disability.

#### 2.2.4 Who will take informed consent?

Written informed consent will be taken by the researchers working within the project team (MZ and JR) in a group meeting lasting up to one hour in length.

### 3.0 Interventions

#### 3.1 Intervention description

The brief problem-solving intervention involves delivery of a well-established social problem-solving theory[29-31]. MZ will deliver the intervention in groups of up to five prisoners in a workshop lasting up to 1.5 hours. The delivery of the intervention will involve (i) watching a digital animation which demonstrates the skills, (ii) completion of a series of workbooks that follow a six-step problem solving model (iii) receiving a demonstration of the problem-solving skills. The model comprises of: Step one: is there a problem?; Step two: describe the problem; Step three: getting information; Step four: think of options; Step five: choose an option; and Step six: make a plan. Because the intervention will be delivered in small groups, the overall duration at each prison site will be up to 1.5 weeks.

The intervention materials have been adapted through prior consultation with up to 50 prison staff and prisoners through co-production workshops and earlier work conducted in the UK and in Poland[26]. All participants regardless of their assignment will also receive their usual care. Usual care comprises of treatment provided by prison staff and medical staff using normal practices and use of medication where appropriate. MZ is a qualified CBT psychotherapist and has received specialist training from AP who devised and developed the intervention in the UK.

#### 3.2 Criteria for discontinuing or modifying allocated interventions

Discontinuation of the intervention delivery can be at the participants' request or by the request of prison staff. Reasons for discontinuation could include (but are not limited to) concerns about risk relating to the prison security or the wellbeing of the participant.

#### 3.3 Strategies to improve adherence to intervention

A shared web-based secure database containing the study identification number, date of randomization, intervention allocation and date of post measurement will be used to support adherence to the intervention. The study identification number will align with each participants questionnaire pack. A small incentive of a notebook and calendar will be offered to participants as a token of appreciation.

**4.0 Outcomes**

The primary outcomes of interest in the feasibility study are recruitment, retention, acceptability and engagement. Data will also be collected on the following measures at baseline and 6 weeks post randomization: depression, general mental health and well-being and coping strategies.

Measurement of depression: The PHQ-9 has shown diagnostic validity in a study of 3,000 adult patients. Each item is rated on a scale of 0 to 3, giving a maximum score of 27. Cut-off scores are used to label depression severity as: 0 to 4, minimal depression; 5 to 9, mild depression; 10 to 14, moderate depression; 15 to 19, moderately severe depression; 20 to 27, severe depression.

Measurement of general mental health and well-being: The General Health Questionnaire – 28 (GHQ-28) is self-report screening measure used to detect possible psychological/psychiatric disorder. The GHQ-28 identifies two main concerns: (1) the inability to carry out normal functions; and (2) the appearance of new and distressing phenomena. The GHQ-28 requests participants to indicate how their health in general has been over the past few weeks, using behavioural items with a 4-point scale indicating the following frequencies of experience: "not at all", "no more than usual", "rather more than usual" and "much more than usual". The scoring system applied in this study was the same as the original scoring system33, the Likert scale 0, 1, 2, 3. The minimum score for the 28 version is 0, and the maximum is 84. Higher GHQ-28 scores indicate higher levels of distress. Goldberg suggests that participants with total scores of 23 or below should be classified as non-psychiatric, while participants with scores > 24 may be classified as psychiatric, but this score is not an absolute cut-off 35.

Measurement of coping strategies: Each item is rated on a scale of 1-4, using 15 sub-scale scores and four statements rated from 'I don't usually do this at all' to 'I usually do this a lot'. Cronbach's alpha for the 15 scales of COPE ranged from .37 to .93. Except for mental disengagement, the remainder of the alphas were all above .59, with the majority above .70. The average alpha was .79.

The study team may also access the participants medical and prison records, and this data may be looked at by the research members during the study.  All data will remain confidential.

#### 4.1 Participant timeline

Table 1 shows the overall trial design with recruitment and measurement time points.

| | STUDY PERIOD June – October 2021 | | | |
|---|---|---|---|---|
| **TIMEPOINT**** | **Enrolment** | **Allocation** | **Intervention delivery** | **6 –weeks post randomization** |
| | **$-t_1$** | **0** | **$t_1$** | |
| **ENROLMENT:** | | | | |
| **Eligibility screen** | X | | | |
| **Information sheet/privacy notice** | X | | | |
| ***[Informed consent]*** | X | | | |
| **Allocation** | | X | | |
| **INTERVENTIONS:** | | | | |
| ***[Problem solving intervention plus usual care]*** | | | X | |
| ***[usual care only]*** | | | X | |
| **ASSESSMENTS:** | | | | |
| ***Demographic questionnaire*** | X | | | |
| ***PHQ-9, GHQ-28, COPE*** | X | | | X |
| ***Acceptability questionnaire*** | | | | X |

#### 4.2 Sample size

The sample size calculations are based on estimating recruitment and attrition rates and standard deviation of the primary outcome measure.  We will randomise 100 participants which will estimation of recruitment (50%) and follow up rates (80%) to be estimated within a 7% and 8% margin of error[32] . A feasibility study of 80 measured subjects will provide robust estimates of standard deviation of the outcome measure in this population to inform the sample size calculation for the subsequent larger definitive fully powered trial[33].

#### 4.3 Recruitment

All participants residing on the therapeutic wing of each prison will be approached by the research team MZ and JR to see if they are willing to participate in the study. The research team will provide information about the study and seek written informed consent using a two-staged approach. Stage one will include an initial introduction to the study, anyone wishing not to take part in the study can decline at this point. In stage two, participants who are interested in finding out more about the study will be invited to attend a group meeting for up to one hour with up to five other prisoners. The meeting will provide an opportunity for participants to hear about the study in detail and ask any questions. At this meeting participants will have the right to decline to and will be able to leave the meeting if they do not want to take part in the study.

Those participants willing to take part will be read aloud the information sheet and relevant privacy notice and consent form. Participants will be encouraged to ask questions about the research project and will then be asked to complete the paperwork and baseline measurements. Participants will not be approached if advised by prison staff that they pose a risk to the researchers. Any participants that come to the attention of the research team as ‘unwell’ either physically or mentally will be referred to the prison doctor or psychologist as per the protocol of the prison. After an independent examination and assessment of the participant, the prison doctor and/or psychologist will recommend whether the participant is well enough to continue participation in the study. Anyone deemed unwell will be removed from the study.

#### 4.4 Assignment of interventions: allocation

##### 4.4.1 Sequence generation

Participants will be randomized by the York Trials Unit Randomization Service at the University of York. This web-based randomization process will randomize patients to one of the two arms of the trial based on a computer-generated code. The information will be stored on a secure server and access to the sequence will be confined to the Trial Manager. Allocation to the trial arms will be in the ratio of 1:1. The Trial Manager will access the treatment allocation for each patient by remote internet-based randomization. The group allocation will be disclosed to the Trial Manager after baseline data has been collected for each participant. The allocation outcome will be entered into the secure shared database so that all members of the research team can view the allocation.

#### 4.5Assignment of interventions: blinding

##### 4.5.1 Who will be blinded?

The pragmatic trial design does not allow us to blind participants, facilitators nor prison staff to the intervention. AP at the University of York will be blind to the outcome assessment data and the allocation of the individuals will not be revealed until all statistical analyses are complete.

#### 4.6 Data collection and management

##### 4.6.1 Plans for assessment and collection of outcomes

The data collection will include four questionnaires collected at two time points (baseline and 6 weeks post-randomization). The data collection procedures and questionnaires will include the following elements:

(i) Check eligibility criteria (first meeting with participant and on the advice of the prison staff). Research team explain the study and request informed consent.
(ii) Gather baseline demographic information (using a specifically devised questionnaire)
(iii) Gather baseline mental health measurement of depression using PHQ-9[34]
(iv) Gather baseline general mental health and Wellbeing using GHQ-28[35]
(v) Gather baseline coping mechanisms and dealing with problems using the COPE[36]

#### 4.7 Data management

All electronic data will be stored for a minimum of 5 years after the end of final analysis of the study. Anonymous data will be entered by JR and will be stored on the shared electronic database giving access to all members of the research team in York and Poland. All paper records will be stored in a secure storage facility in Poland. Personal identifiable paper records will be stored separately from anonymised paper records and will remain in Poland. As a backup, all electronic data records will be stored also on a password protected server within the Health Sciences Department at the University of York.

#### 4.8 Confidentiality

Participants will complete a written informed consent form and will receive a separate privacy notice from the coordinating Centre (University of York in the UK) and the collaborators (The Academy of Justice in Poland). All data will remain confidential.

**5.0 Statistical methods**

The flow of participants through the trial will be detailed in a CONSORT flow diagram. The number of people screened, randomly assigned, receiving the intervention, completing the study protocol, and providing outcome data will be summarized overall and by randomized group. The number of individuals withdrawing from the intervention and/or the trial and any reasons for withdrawal will be summarized by group. This feasibility study is not powered to formally assess the size of the treatment effect, rather to estimate the recruitment rate. However, the totality of the data collected will be used to assess the feasibility of a definitive large RCT; recruitment rate being the driver of the feasibility study design on the basis that unless a reasonable recruitment rate can be achieved no formal trial would be possible. The recruitment rate will be estimated based on data collected and a 95% confidence interval determined for this measure. The outcome measures (PHQ-9; GHQ-28; COPE) will be reported descriptively and completion rates compared between study groups. All outcomes will be summarized descriptively using mean, SD, median, 25th and 75th percentiles for continuous outcomes and the number of events and percentages for categorical data. To quantify the acceptability of the intervention the number of sessions attended, and acceptability responses will also be summarized.

**5.2 Methods in analysis to handle protocol non-adherence and any statistical methods to handle missing data**

Missing data will be summarized for each outcome overall and by study group.

**6.0 Ethical issues**

We do not anticipate any major ethical issues. Where participation in the trial is felt to be detrimental to health and wellbeing, we will not make an approach to participate. Participants will not be denied any form of care that is currently available in the Polish prison system. The trial does not involve new medicinal products or any invasive/potentially harmful procedures and is therefore considered low risk for participants. All participants will receive usual care, and therefore no treatment will be withheld by participating in this trial. The research team will be guided by the prison staff as to any risk posed by approaching individuals to take part in the trial. Written informed consent will be obtained from every participant. Permission to conduct the study has been granted by the Director of each prison and the Prison staff at each Polish prison, The Academy of Justice in Warsaw and the Health Sciences Department at the University of York.

#### 6.1 Oversight and monitoring

##### 6.1.1 Composition of the coordinating Centre and trial steering committee

The coordinating Centre at the University of York includes the study PI and the York Trials Unit. The coordinating Centre will be responsible for the management and oversight of the trial and will provide day to day support for those conducting the research in Poland. The coordinating Centre will provide the shared electronic databases to ensure integrity to the randomization process and the database template for entry of the data at baseline and post assessments. The coordinating Centre will conduct the randomization process and allocation of participants to the trial. The research team in Poland are responsible for the recruitment, informed consent baseline and post-test intervention data collection and delivery of the intervention. The research team in Poland will also enter the required demographic information for the randomization process and enter the collected data into the shared electronic database. The research team in Poland will retain and store in secure provision all papers copies of the data with consent forms stored separately from the data.

##### 6.1.2 Composition of the data monitoring committee, its role and reporting structure

The trial will not require a data monitoring committee as the numbers in the trial will be small enough to be monitored by the research team at York University. The project has been reviewed by the RODO data protection specialist at The Academy of Justice in Warsaw, Poland and each prison site has an information data officer who can be contacted. As part of our information sheets and privacy notice we have provided participants with access to complain independently and report any concerns about data management or breech of data confidentiality in our study paperwork for the UK and in Poland.

##### 6.1.3 Adverse event reporting and harms

This study is non-CTIMP (Clinical Trial of an Investigational Medicinal Product) and is therefore not subject to any additional restrictions. Decisions regarding prescription of any medications will made by the participant in conjunction with their usual care, participation in the study will have no bearing on this process. This study will record details of any Serious Adverse Events (SAEs) that are required to be reported to the Governance Committee at the University of York, The Academy of Justice in Poland and the prison Director at each prison site. Under the terms of the Standard Operating

Procedures for RECs [49]. An SAE is defined as a 'related'* and 'unexpected'** untoward occurrence that:

(a) Results in death.

(b) Is life threatening.

(c) Requires hospitalization or prolongation of existing hospitalization.

(d) Results in persistent or significant disability or incapacity.

(e) Consists of a congenital anomaly or birth defect; or

(f) Is otherwise considered medically significant by the investigator.

* 'related' is defined as: resulting from the administration of any research procedures.

** 'unexpected' is defined as: a type of event not listed in the protocol as an expected occurrence.

In the context of the current study, an occurrence of the type listed in (a) to (f) above will be reported as an SAE only if:

a) It is suspected to be related to an aspect of the research procedures (e.g. completion of follow-up questionnaires, participation in delivery of the intervention).

Or

b) It is an unexpected occurrence. Hospitalization's, disabling / incapacitating / life-threatening conditions and deaths are expected in the study population due to the age of the cohort, they will therefore only be reported as SAEs if they appear to be related to an aspect of taking part in the study.

Polish colleagues will inform the research team in York who will decide if the event should be reported to the York Research Governance Committee as an SAE. Related and unexpected SAEs will be reported to the main REC within 15 days of the PI becoming aware of the event. A SAE Form will be completed, and a copy stored in the participant's records.

**7.0 Plans for communicating important protocol amendments to relevant parties (e.g. trial participants, ethical committees)**

Changes to the protocol (e.g., changes to eligibility criteria, outcomes, analyses) will be notified to relevant parties (e.g., investigators, governance committees, trial participants, trial registries, journals, regulators). An updated version number of the protocol will be published to reflect any changes that are made.

#### 7.1 Dissemination plans

We will publish a paper relating to this trial that will include (as a minimum) the results of the feasibility trial.  We will produce a short summary of the results that can be distributed to all trial in the form of a poster. Finally, we will aim to ensure coverage of our findings in the wider media by issuing a press release. Use of social media (twitter) and the project webpages (https://www.york.ac.uk/healthsciences/research/mental-health/projects/international-adaptation-poland/) will help to disseminate information to the wider public.

### 8.0 Trial status

Protocol v2 18/05/2021

Date of first participant recruitment: Not yet started

### 8.1 Declarations

### 8.2 Funding

Funding is supported through the Centre for Future Research at the University of York, Funding in kind is supported through the research team and the provision of incentives to the participants for taking part in the study at The Academy of Justice in Warsaw.
